# Heart Failure Prediction: An Explainable Cross-Validated Comparison of Several Machine Learning Models

**DOI:** 10.1101/2025.10.21.25338436

**Authors:** Md Sumon Ali, Towhidul Islam

## Abstract

Heart disease is the leading cause of death worldwide, contributing to millions of fatalities each year. Early detection and accurate risk prediction are therefore critical for timely intervention and improved patient outcomes. In this study, seven different Machine Learning (ML) prediction models were tested on the collections of heart disease datasets. The data was divided, with 80% used to train the models and 20% saved to test them later. The settings for the models were carefully adjusted using a method that checks their performance multiple times to find the best ones. The models were then evaluated on the unseen test data. Their performance was measured using several metrics (Accuracy, Recall, Precision, PR-AUC and ROC-AUC), and their results were further examined using a confusion matrix. In addition to traditional evaluation metrics, SHapley Additive exPlanations (SHAP) analysis was employed to interpret the contribution of each feature to the model’s predictions. It was observed that the Multi-Layer Perceptron (MLP) achieved the highest performance on both datasets, demonstrating strong predictive capability while remaining interpretable through the integration of SHAP. This study shows that modern ML models can be very good at predicting heart disease risk, and provide explainable performance. This study provides an effective approach for predicting heart disease risk with an explainable model to help doctors choose the best tool.

## 1 Introduction

One of the most important organs in the human body is the heart. Its duties include pumping oxygenated blood via arteries to every other part of the body and receiving deoxygenated blood from the body through blood vessels, which also purify and transport metabolic waste. The term Cardiovascular Disease (CVD) is generally referred to as the condition affecting the heart or blood vessels[1]. The world health organization estimates that 17.9 million people worldwide are at risk of dying from CVDs, which are the leading cause of death worldwide[2]. Early detection and treatment of heart disease are getting more challenging due to population growth. The goal is to build Machine Learning (ML) algorithms than can predict the likelihood of heart disease based on various health metrics and features [3–6]. With application in a variety industries, including healthcare, ML is one of the branches of Artificial Intelligence (AI) that is expanding fastest. Because ML is an intelligent tool for analyzing data, and the medical profession is abundant with data, it offers a lot of value in the healthcare field [7].

A healthy heart is conducive to a healthy life. The majority of people experience stress, anxiety, depression, and a variety of other illnesses as a result of their hectic lifestyles. These major factors are causing people to become ill and experience severe illnesses[8]. In the modern world, CVD is becoming a bigger concern. Sudden arrest of Aldous can lead to serious illness such as brain damage, nervous system disorder and even death. Because of this, heart failure condition can be identified early rather than later [9]. Current strategies for predicting risk are only modestly successful, likely because they are derived from traditional statistical analysis methods that fail to capture prognostic information in large datasets containing multi-dimensional interactions [10]. Furthermore, most existing approaches lack an explainable framework, which limits their interpretability and clinical trust [11, 12]. However, they can be insufficient due to their limited ability to handle complex patterns in data, often resulting in lower accuracy or generalizability [13].

By applying and analyzing a range of modern ML algorithms across multiple datasets, including Random Forest (RF), Logistic Regression (LR), Support Vector Classifier (SVC), k-Nearest Neighbors (KNN), Decision Tree (DT), XGBoost, and Multilayer Perceptron (MLP), and by fine tuning their hyperparameters using techniques such as cross-validation and grid search to ensure reliable performance, this research aims to enhance predictive models for heart disease risk. In addition, SHapley Additive exPlanations (SHAP) is employed to interpret the contribution of each feature to the model’s predictions, providing transparency and clinical insight. The novel aspects of this work include improving computational efficiency, increasing forecast accuracy, and incorporating explainable predictions to support trustworthy clinical decision-making.

The main contributions of this work can be summarized as follows:

- Development and evaluation of multiple modern ML models, including RF, LR, SVC, KNN, DT, XGBoost, and MLP, for predicting heart disease risk across multiple datasets with fine-tuned hyperparameters using cross-validation and grid search.
- Integration of SHAP to provide interpretable and transparent predictions, allowing identification of the most influential features and enhancing clinical trust in the models.
- Proposal of a robust and computationally efficient predictive framework that improves forecast accuracy while addressing limitations of traditional statistical models, such as handling complex patterns and multi-dimensional interactions in medical data.

The rest of the paper is organized as: section 2 describes the related work and section 3 outlines the methodology, while section 4 describes the results, which are discussed in section 5, finally, section 6 concludes the paper and puts forward some future directions.

## 2 Literature Review

In recent years, the application of ML algorithms in medical research has gained significant attention. Advances in medical science have been largely driven by the proliferation of information and communication technologies [14]. The large-scale collection of medical data enabled by smart healthcare technologies has facilitated the development of numerous ML and Deep Learning (DL) techniques for improved diagnostic and prognostic outcomes [15].

Cardiovascular diseases remain a major health concern, with modern lifestyle factors, such as increased exposure to dietary pesticides and sedentary behavior, contributing to a rising risk of heart failure [16]. Many studies have employed ML algorithms including SVC, RF, DT, XGBoost, KNN, and Neural Networks (NN) to improve the accuracy of heart disease prediction (Table 1). These approaches have demonstrated enhanced predictive performance. However, several limitations persist. Most studies rely on a single dataset, suffer from demographic biases, and face scalability challenges, indicating a need for more generalized and robust predictive models.

**Table 1:** Overview of Machine Learning Approaches, Evaluation Metrics, and Datasets for Heart Disease Diagnosis.

| Year | ML Techniques | Evaluation Measures | Data Sets | Limitation |
| --- | --- | --- | --- | --- |
| 2024[18] | RF,<br>LR,<br>Naïve Bayes,<br>SVM, DT,<br>KNN, XGBoost,<br>NN | Accuracy | Heart disease dataset | The study's performance heavily depends on the dataset's size and quality also lack of explainability. |
| 2022[19] | LR,<br>KNN, DT,<br>XGBoost (Ensemble classifier) | Accuracy,<br>TP Rate,<br>FP Rate,<br>Precision,<br>ROC Curve,<br>Kappa Statistics | Cardiovascular dataset | The study suggests that scalability and accuracy could be further improved. |
| 2024[20] | Naïve Bayes Classifier | GPS,<br>AUC of the ROC | Spanish Delta@ database | The model does not account for individual health conditions or personal habits of workers. |
| 2023[5] | LR,<br>KNN,<br>SVM,<br>RF,<br>Gradient Boosting | ROC_AUC,<br>Precision,<br>Recall,<br>Accuracy | UCI Heart Disease Dataset | The paper's limitation lies in its reliance on traditional ML models. |
| 2020[10] | Boosted Decision Tree (BDT) | AUC,<br>ROC,<br>C-statistic | UCSD,<br>UCSF,<br>BIOSTAT-CHF cohorts | The model was derived from a single center, which may introduce demographic selection bias. |
| 2019[21] | Hybrid RF with Linear Model (HRFLM) | Accuracy, Precision,<br>F-measure,<br>Sensitivity,<br>Specificity | Cleveland heart disease dataset | The study is limited by using a single dataset. |
| 2021[22] | KNN,<br>DT,<br>RF,<br>LR,<br>AdaBoostM1,<br>MLP | Accuracy,<br>Sensitivity, Specificity,<br>Kappa,<br>MCC,<br>F-Measure | Heart disease dataset | The study acknowledges that the dataset size was not large enough. |

Another important limitation of prior work is the lack of explainable frameworks. Although these models achieve high predictive accuracy, they often operate as “black boxes”, offering little insight into the contribution of individual features to predictions. The use of SHAP can address this limitation by providing both global and local interpretability of model predictions [11, 12, 17]. Despite its potential, very few studies have applied SHAP to understand feature importance and enhance transparency in heart failure risk prediction, highlighting an opportunity for integrating explainable ML in clinical decision support.

In this research, two distinct datasets are used to address these limitations. This research also apply a uniform ML model to improve the robustness and generalizability of the findings. This method aims to solve the previous challenges by leveraging different datasets to enhance predictive accuracy and mitigate model biases, thus contributing to more reliable frameworks for heart disease prediction.

## 3 Methodology

### 3.1 Dataset Analysis

For every dataset, at first we need to process the data which is shown is Fig. 1. The dataset preprocessing steps were performed to prepare the data for training the model and evaluation.

**Fig. 1:**
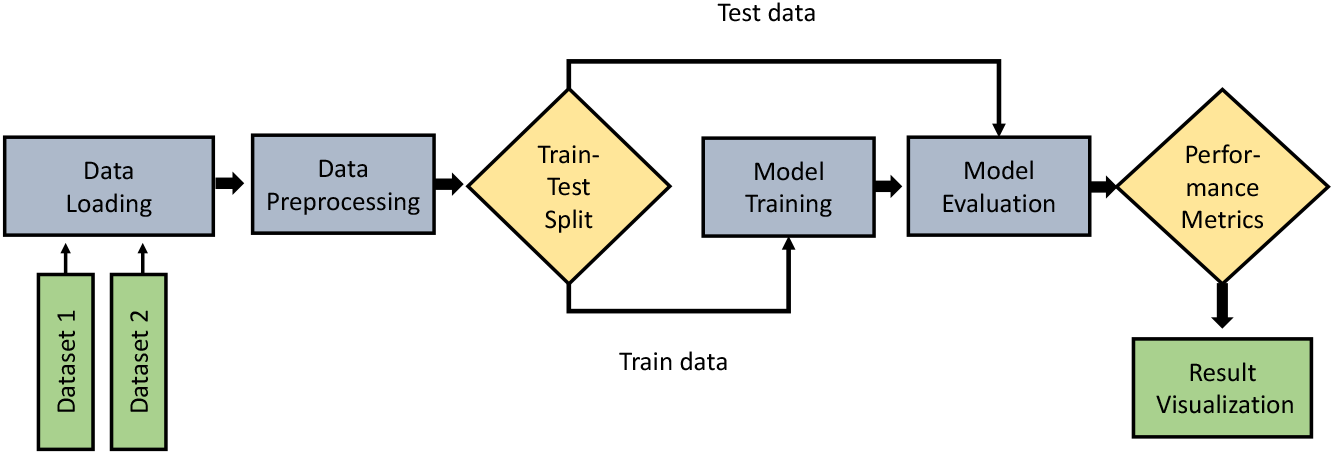
Proposed method for heart failure prediction.

#### 3.1.1 Dataset 1: Heart Failure Prediction Dataset

With an estimated 17.9 million fatalities annually, or 31% of all deaths worldwide, CVDs are the leading cause of mortality worldwide. This dataset features 12(Age, Sex, ChestPainType, RestingBP, Cholesterol, FastingBS, RestingECG, MaxHR, ExceriseAngina, Oldpeak, STSlope and HeartDiease) variables that can be used to predict a potential heart illness.The output label of this dataset is HeartDisease. There are 5 categorical, 5 Numerical and 2 Binary feature in the dataset. The dataset contains 55.3% (HeartDiease=1) and 44.7% (HeartDiease=0), which looks somehow balanced. Based on the domain knowledge, features such as ChestPainType, ExerciseAngina, and Oldpeak have strong predictors of heart disease. The dataset apply on the automated feature engineering techniques, but it lacks manual feature engineering derived from domain expertise. This dataset is collected from Kaggle[23].

There are multiple steps involved to prepare the Heart Failure Prediction Dataset. At first, numerical and categorical features are isolated. Numerical features that contain zero values are replaced by using median imputation grouped by specific columns. Negative values are set to NaN, then filled with median values using an imputer. Categorical features were converted to numerical via one-hot encoded[24]. Then the datasets were split into train and test (80-20) ratio, followed by feature scaling through standard scalar. Fig. 2 shows the correlation matrix for Heart Failure Prediction Dataset. This heat map matrix is useful for identifying strong correlations. This relation indicates redundant information or interactions. Accuracy, Recall, ROC-AUC, and PR-AUC are the metrics to evaluate the datasets. Pandas, StandardScalar, and Matplotlib are the frameworks used for this datasets.

**Fig. 2:**
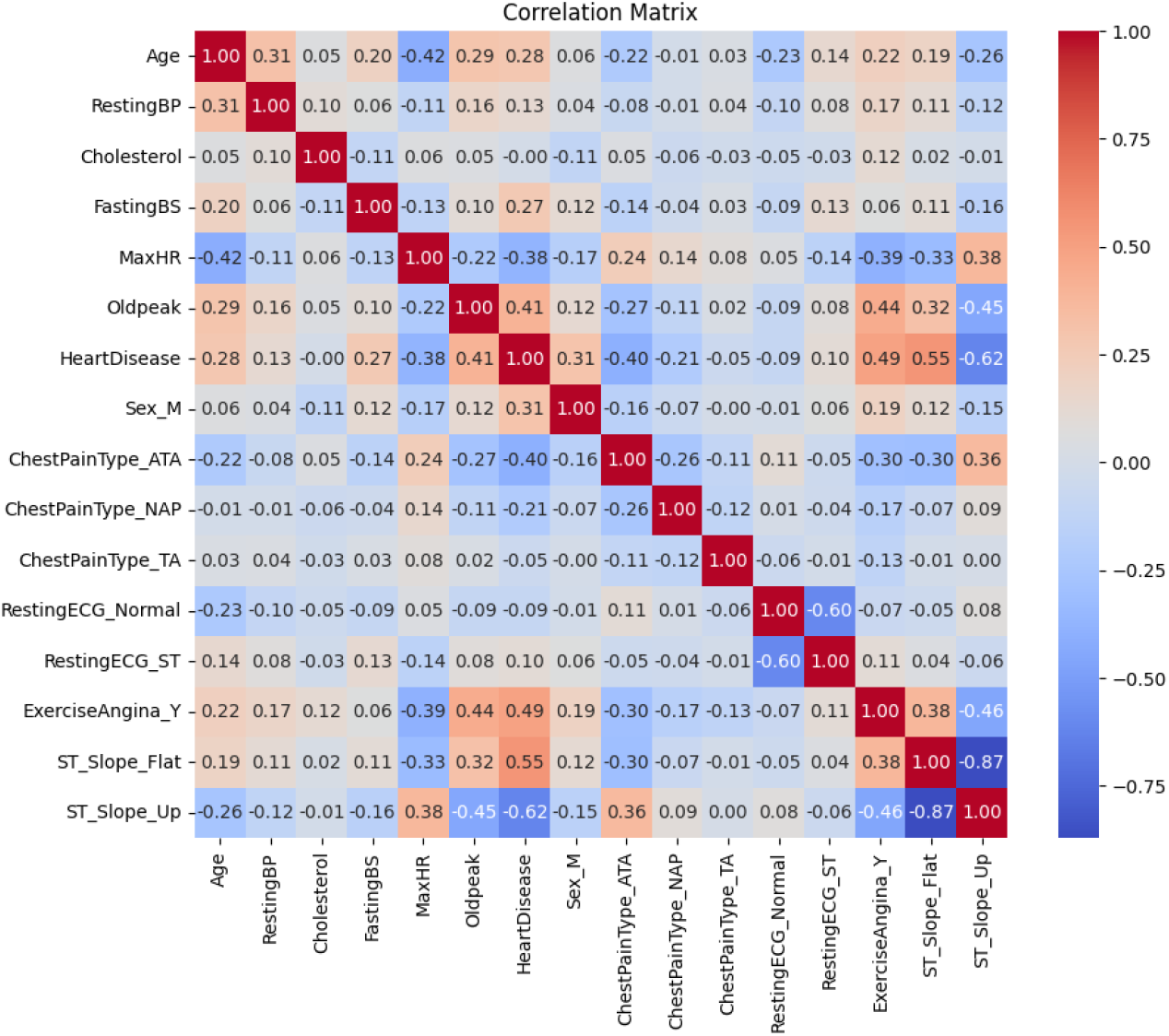
Correlation matrix of Dataset 1.

#### 3.1.2 Dataset 2: Heart Attack Analysis and Prediction Dataset

This dataset was collected from kaggle[25]. This dataset has 14 (age, sex, chest pain type, resting blood pressure, cholesterol, fasting blood sugar, resting electrocardiogram, maximum heart rate, exercise induced angina, oldpeak, slope of the peak exercise ST segment, number of major vessels, thalassemia, and output) features. There are 5 numerical features, 4 categorical (binary) features, and 5 categorical (non-binary) feature in the dataset. One hot enconding technique converts categorical to numerical value. Chest pain type, thalachh, oldpeak, and caa are the expected important features of the datasets. The output label is output, which is 0 and 1. The dataset is not significantly imbalanced where output=1 is 54.46% and output=0 is 45.54%.

This dataset calculated the Interquartile Range (IQR) for each column to define outlier boundaries[26]. Then a correlation matrix is computed, helping to understand the relationships between variables, which is shown in Fig. 3. The dataset is split into an 80-20 ratio. Standard scaling is applied to ensure that all features are on a similar scale[27]. Accuracy, Recall, ROC-AUC, and PR-AUC are the metrics to evaluate the datasets. Pandas, StandardScalar, and Matplotlib are the frameworks used for this datasets.

**Fig. 3:**
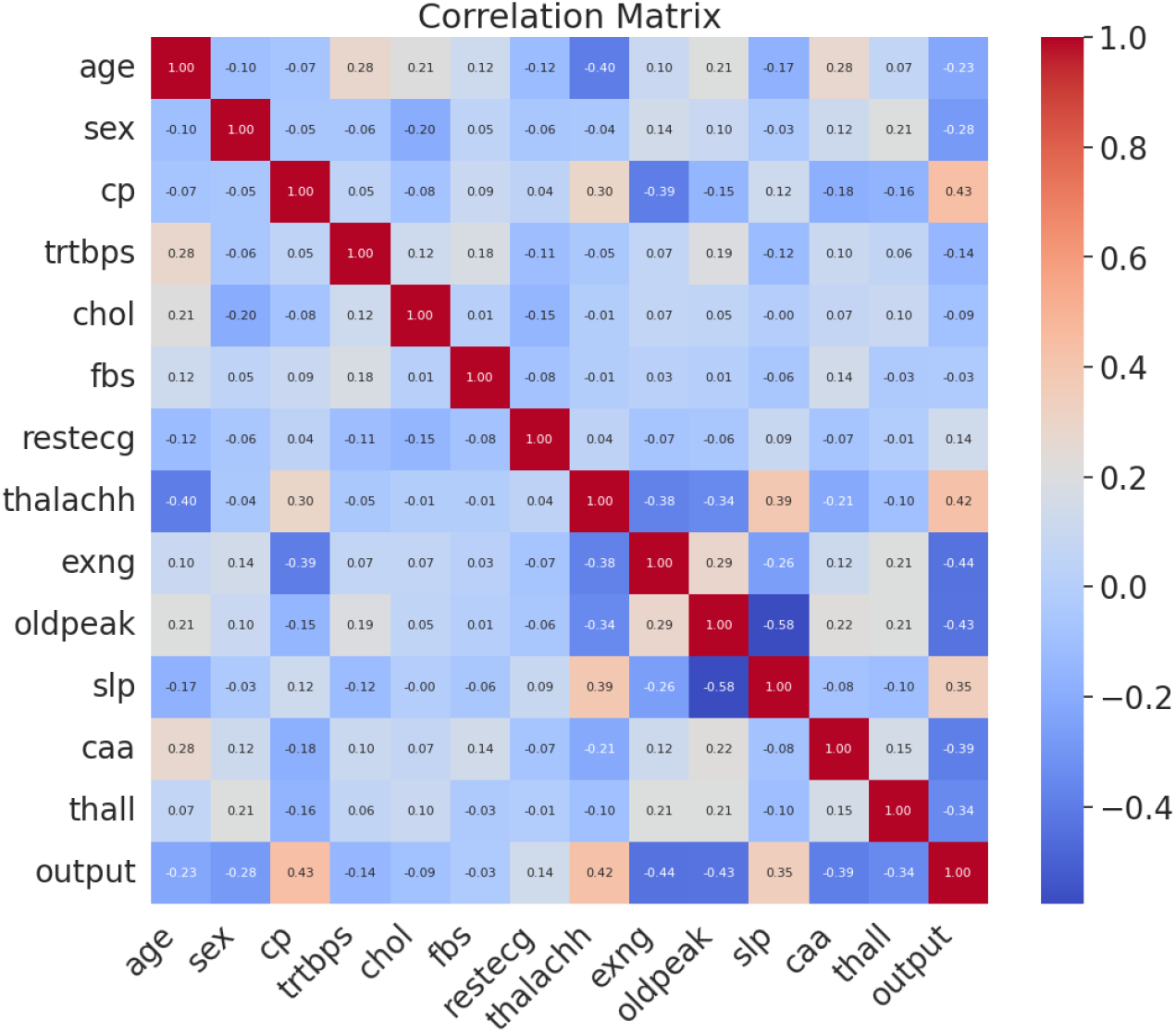
Heat map for Heart Attack Analysis and Prediction Dataset 2.

### 3.2 Model Selection

A diverse pool of ML modes was selected to investigate their performance using different datasets which is shwon in Fig 4. The datasets were split using train-test-split with an 80-20 split ratio. The model selection process involved using Stratified K-Fold Cross-Validation with 5 splits. This method ensures the robustness of the methodology.

**Fig. 4:**
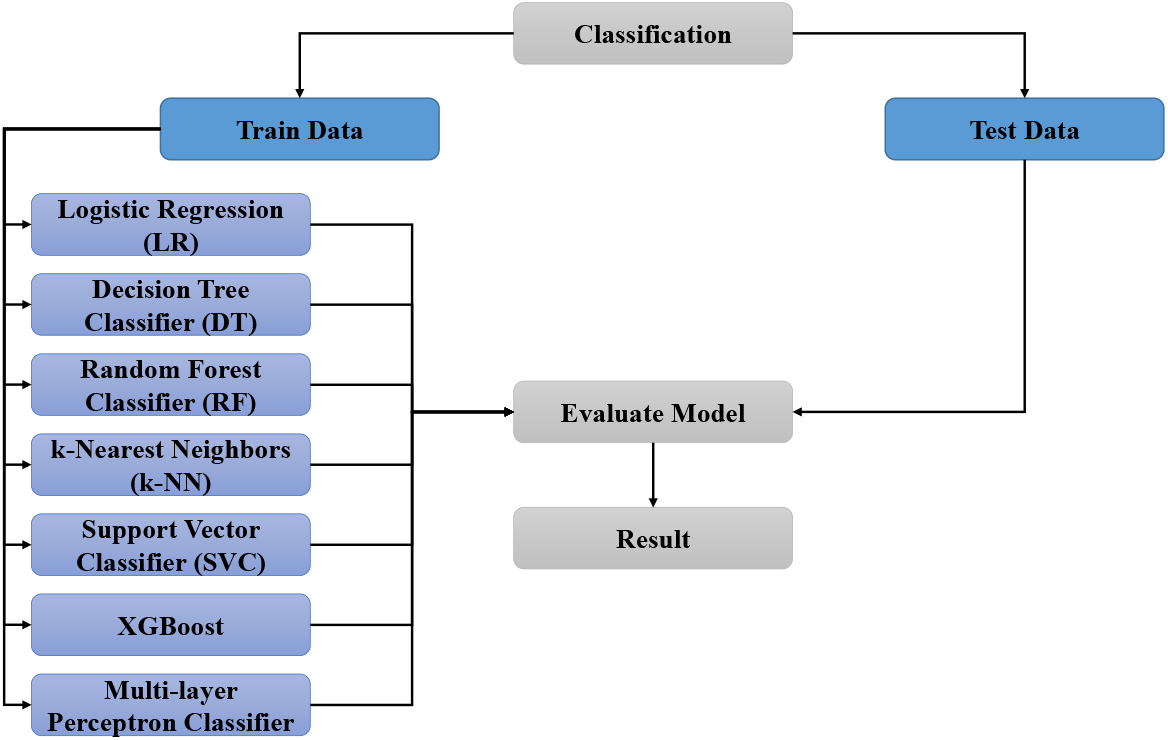
Machine Learning Algorithms Pool.

#### 3.2.1 Hyperparameter Tuning

To find out the optimal model performance, hyperparameter tuning was conducted using GridSearchCV[28]. In this case, we consider a 5-fold cross-validation strategy for each model in the pool. Different types of hyperparameters were tuned for each model, including:

##### Logistic Regression (LR)

Penalties l1(lasso), l2(Ridge), Elastic, and none were applied to the logistic regression to prevent overfitting. Logistic regression also used solvers (lbfgs, liblinear, saga) to optimize the model which is shown in Table 2. In the case of logistic regression, the total hyperparameter combination is 480.

**Table 2:**
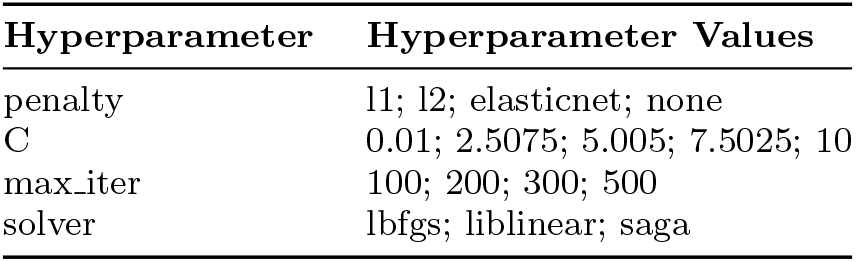
Hyperparameters and their values for LR.

##### K-Nearest Neighbors (KNN)

To find out the best performance of the KNN model, a hyperparameter tuning process was introduced with a grid search technique. The number of neighbors defined in the case of KNN was 3, 5, 7, 10, and 15. Two weight parameters (uniform, distance) control the weight assignment to neighbors when making predictions. The algorithm parameter (auto, ball tree, kd tree, and brute) specifies the algorithm used to compute the nearest neighbors. Finally, leaf size (10, 20, 30, 40, 50) affects the speed of the Ball Tree and KD Tree algorithms, particularly during the search phase which is shown in Table 3. KNN contains a 400 hyperparameters combination.

**Table 3:** Hyperparameters and their values for KNN model.

| Hyperparameter | Hyperparameter Values |
| --- | --- |
| n_neighbors | 3; 5; 7; 10; 15 |
| weights | uniform; distance |
| algorithm | auto; ball_tree; kd_tree; brute |
| leaf_size | 10; 20; 30; 40; 50 |

##### Decision Tree Classifier (DT)

The hyperparameters criterion determines the function used to measure the quality of a split. In Criterion two options were considered. The Gini impurity metric used to evaluate splits and entropy employs the information gain[29]. The maximum depth parameter limits the depth of the decision tree. The values explored ranged from 10 to 50 in increments of 10, with the additional option None to allow unrestricted tree depth which is shown in Table 4. DT contains a combination 216 hyperparameters.

**Table 4:** Hyperparameters and their values for DT model.

| Hyperparameter | Hyperparameter Values |
| --- | --- |
| criterion | gini; entropy |
| max_depth | 10; 20; 30; 40; 50; None |
| min_samples_split | 2; 5; 10 |
| min_samples_leaf | 1; 2; 4 |

##### Random Forest Classifier (RF)

The number of estimators in RF defines the number of decision trees. Values ranging from 50 to 200 were used in increments of approximately 50. Maximum depth controls the depth of each individual decision tree. The maximum depth values explored ranged from 10 to 50, with increments of 10. Minimum sample split specifies the minimum number of samples required to split an internal node, and minimum sample per leaf defines the number of samples required to form a leaf node which is shown in Table 5. RF contains a combination of 432 hyperparameters.

**Table 5:** Hyperparameters and their values for RF model.

| Hyperparameter | Hyperparameter Values |
| --- | --- |
| n_estimators | 50; 100; 150; 200 |
| max_depth | 10; 20; 30; 40; 50; None |
| min_samples_split | 2; 5; 10 |
| min_samples_leaf | 1; 2; 4 |

##### Support Vector Classifier (SVC)

In case of SVC, the kernel function defines how the data is transformed before classification. Four types of kernels were tested: linear, poly, rbf, and sigmoid. The regularization parameter was tested to control the trade-off between maximizing the margin and minimizing classification errors. The kernel coefficient (gamma) determines the influence of individual training samples on the decision boundary which is shown in Table 6. SVC contains 240 hyperparameters combination.

**Table 6:** Hyperparameters and their values for SVC model.

| Hyperparameter | Hyperparameter Values |
| --- | --- |
| kernel | linear; poly; rbf; sigmoid |
| C | 0.01; 2.5075; 5.005; 7.5025; 10 |
| gamma | scale; auto |
| degree | 2; 3; 4 |

##### Extreme Gradient Boosting Classifier (XGBoost)

The gradient boosting approach, which is linked to the decision tree technique, has been used in a variety of fields recently. In many regression and classification situations, XGBoost works better when paired with other techniques [30]. The latest version of the gradient boost technique is called XGBOOST (Extreme Gradient Boosting)[31]. Number of estimators (50-200), maximum depth (3,5,7), learning rate (0.01, 0.2, 3), subsample (0.8, 0.9, 1.0), and column subsample (0.8, 0.9, 1.0) are the hyperparameters in XGBoost model. By conducting a comprehensive grid search over the hyperparameters, the optimal combination was identified to maximize the predictive performance of the XGBoostd model which is shown in Table 7. XGBoosts contains 648 hyperparameters combination.

**Table 7:** Hyperparameters and their values for XGBoost model.

| Hyperparameter | Hyperparameter Values |
| --- | --- |
| n_estimators | 50; 100; 150; 200 |
| max_depth | 3; 5; 7 |
| learning_rate | 0.01; 0.105; 0.2 |
| subsample | 0.8; 0.9; 1.0 |
| colsample_bytree | 0.8; 0.9; 1.0 |

##### Multi-layer Perceptron Classifier (MLP)

The MLP is a well-known Artificial Neural Network (ANN). This robust modeling tool uses data samples with known outcomes to apply a supervised training process[32]. In this study, we show that a computational model based on a multilayer perceptron network with three layers—one single hidden layer, and two hidden layer—can be used to construct such a complicated system to enable the detection of cardiac disease. Relu and tanh activation functions are used to introduce non-linearity into the model which is shown in Table 8. To solve, Adam and SGD were used for weight updates during training. MLP contains a combination of 240 hyperparameters.

**Table 8:** Hyperparameters and their values for MLP model.

| Hyperparameter | Hyperparameter Values |
| --- | --- |
| hidden_layer_sizes | (50,); (100,); (50, 50) |
| activation | relu; tanh |
| solver | adam; sgd |
| alpha | 0.0001; 0.002575; 0.00505; 0.007525; 0.01 |
| learning_rate | constant; adaptive |

Combination of seven ML model framework was established, each optimized through an extensive grid search for hyperparameter tuning. For each model, a combination of core hyperparamters was tested, resulting in a total of 2656 hyperparamter configurations across all models. The search space was extended to include polynomial feature transformations (degrees 1 and 2) to find out non-linear relationships in the datasets. For each dataset, GridSearchCV was used to select the best hyperparameters based on cross-validated accuracy. This approach facilitated a comprehensive search over parameter spaces to identify the most suitable configuration.

To understand the contribution of individual features to model predictions and enhance clinical trust, SHapley Additive exPlanations (SHAP) [11, 12] was applied to each optimized model. SHAP provides a unified framework for quantifying feature importance at both the global and local levels which is shown in Fig. 6, and Fig. 8. This analysis allows identification of the most influential clinical factors for heart disease prediction and ensures that the model’s decisions are interpretable and clinically meaningful.

## 4 Analyze the Results and Find the Best Hyperparameters

Each model’s performance was evaluated using multiple metrics to ensure robustness of the research. The proportion of correctly classified samples was identified by Accuracy. Recall is the ability of the model to identify positive instances correctly. ROC-AUC measured the trade-off between the true positive rate and false positive rate. Finally, the PR-AUC focuses on the precision-recall trade-off, which is especially relevant in imbalance datasets. These metrics were calculated for each fold in the 5-fold cross-validation, and the average performance across fold was reported.

### Dataset 1

Aggregate the cross-validation results (Table 9) by calculating the mean scores for each model, then rank the models based on the PR-AUC metric which is shown in Table 10. Then, select the model with the highest score on the PR-AUC. Finally, found the hyperparameter for the best-performing model.

**Table 9:**
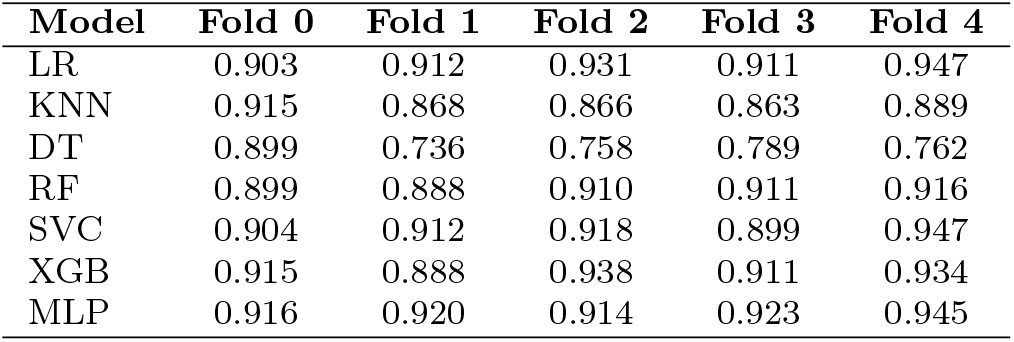
Cross-validation performance (AUC) of different models across five folds for Dataset 1.

**Table 10:** Ranking of models based on mean Precision-Recall AUC across five-fold cross-validation for Dataset 1.

| Rank | Model | Mean PR AUC |
| --- | --- | --- |
| 1 | MLP | 0.924 |
| 2 | LR | 0.921 |
| 3 | XGB | 0.917 |
| 4 | SVC | 0.916 |
| 5 | RF | 0.905 |
| 6 | KNN | 0.880 |
| 7 | DT | 0.789 |

The best parameters for the best model are: `mlp_activation: relu, mlp_alpha`: 0.0001, `mlp_hidden_layer_sizes`: (50, 50), `mlp_learning_rate: constant, ml_solver: sgd, poly_degree:` 1.

### Dataset 2

Aggregate the cross-validation results (Table 11) by calculating the mean scores for each model, then rank the models based on the PR-AUC metric which is shown in Table 12. Then, select the model with the highest score on the PR-AUC. Finally, found the hyperparameters for the best-performing model.

**Table 11:** Precision–Recall AUC values for each model across five-fold cross-validation for Dataset 2.

| Model | Fold 0 | Fold 1 | Fold 2 | Fold 3 | Fold 4 |
| --- | --- | --- | --- | --- | --- |
| LR | 0.947 | 0.859 | 0.926 | 0.894 | 0.910 |
| KNN | 0.900 | 0.799 | 0.821 | 0.894 | 0.879 |
| DT | 0.852 | 0.778 | 0.749 | 0.898 | 0.763 |
| RF | 0.885 | 0.862 | 0.924 | 0.900 | 0.868 |
| SVC | 0.941 | 0.899 | 0.924 | 0.924 | 0.868 |
| XGB | 0.918 | 0.888 | 0.924 | 0.912 | 0.863 |
| MLP | 0.931 | 0.888 | 0.942 | 0.910 | 0.910 |

**Table 12:** Ranking of models based on mean Precision–Recall AUC across five-fold cross-validation for Dataset 2.

| Rank | Model | Mean PR AUC |
| --- | --- | --- |
| 1 | MLP | 0.916 |
| 2 | SVC | 0.911 |
| 3 | LR | 0.907 |
| 4 | XGB | 0.901 |
| 5 | RF | 0.888 |
| 6 | KNN | 0.859 |
| 7 | DT | 0.808 |

The best parameters for the best model are: `mlp_activation:` ‘relu’, `mlp_alpha:` 0.0001, `mlp_hidden_layer_sizes:` (50,), mlp_learning_rate: ‘constant’, `mlp_solver: ‘sgd’`, `poly_degree:`1.

### 4.1 Final Evaluation Using Best hyperparameters

This is the final test for the best hyperparameters. From the full dataset, we created an 80/20 split. The 80% portion was used for training and to fit the scaler, that fitted scaler was then applied to the remaining 20%, which was held out (never used during model development) and used only for final testing.

### 4.2 Analyze the test Result

The performance of all datasets is shown in Table 13. In case of Dataset 1 the overall model performance is strong and acceptable. The model achieved accuracy of 86.41%, which indicating a high overall correctness in predictions. The model is performing quite well in Dataset 2 and acceptable. The accuracy of 83.61% indicate that model is generally effective at predicting heart disease.

**Table 13:** Best Model Performance Across Datasets.

| Dataset | Best Model | Accuracy | Recall | Precision | ROC AUC | PR AUC |
| --- | --- | --- | --- | --- | --- | --- |
| Dataset 1 | MLP | 0.864 | 0.859 | 0.903 | 0.865 | 0.957 |
| Dataset 2 | MLP | 0.836 | 0.812 | 0.864 | 0.837 | 0.906 |

The confusion matrix for all datasets is shown in Fig. 5. For Dataset 1, the model achieves a relatively high number of True Positive (92) and True Negatives (67) but also produces 15 False Negatives and 10 False Positive. This results provide a good balance between sensitivity and specificity. For Datasets 2 False Positives (4) and False Negatives (6) which indicating that the model is more conservative. The model is avoiding Type I errors.

**Fig. 5:**
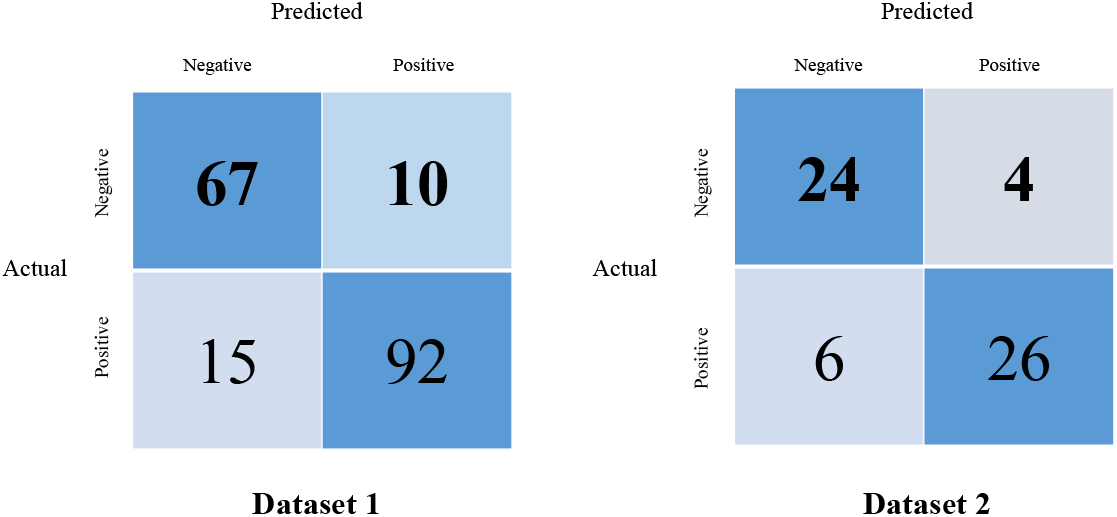
Confusion Matrices for Datasets 1 and Dataset 2.

**Fig. 6:**
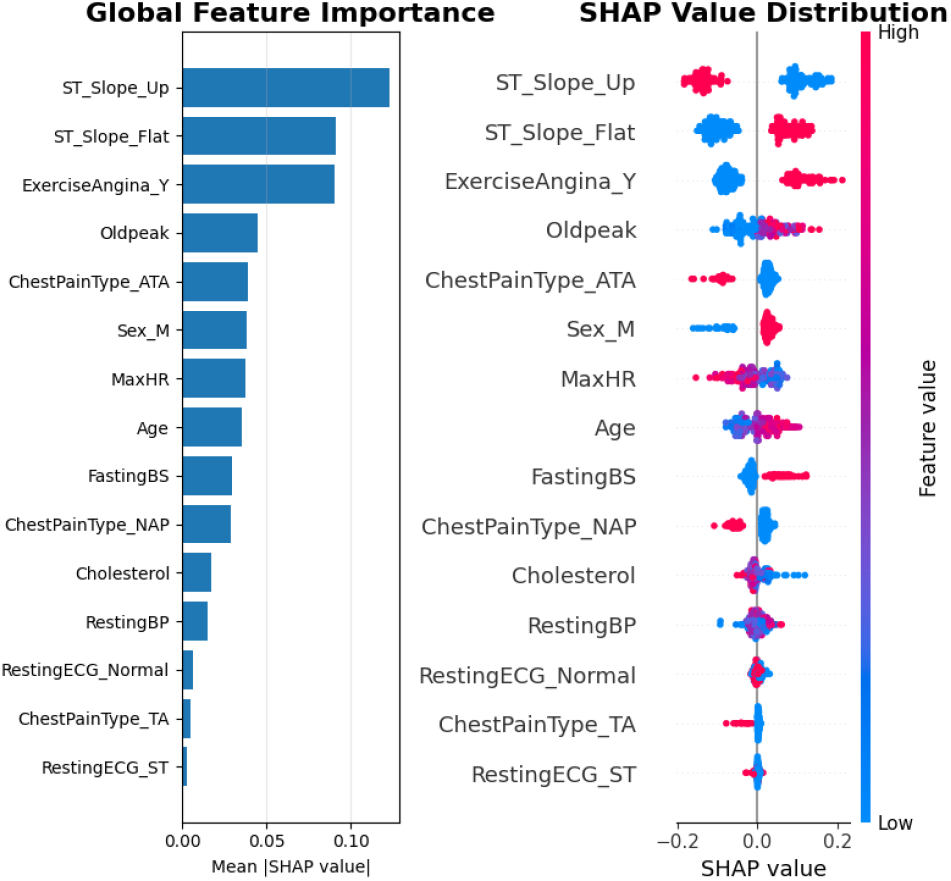
Global Feature Importance and SHAP Value Distribution for Dataset 1.

### 4.3 Model Interpretability Using SHAP

To interpret the predictions of the best-performing models and understand feature contributions, SHAP was applied. SHAP provides both global and local interpretability:

- **Global interpretation:** Identifies which features contribute most on average to model predictions across the dataset, using mean absolute SHAP values. The global feature importance and SHAP value distribution for Dataset 1 and Dataset 2 are shown in Fig. 6 and Fig. 8, respectively.
- **Local interpretation:** Examines individual predictions to show how each feature pushes the prediction towards higher or lower risk. SHAP force plots for sample 0 in Dataset 1 and Dataset 2 are shown in Fig. 7 and Fig. 9, respectively.

**Fig. 7:**
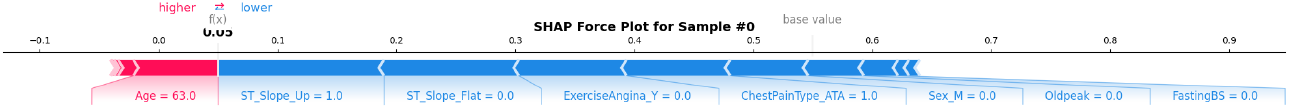
SHAP Force Plot for Sample 0 in Dataset 1.

**Fig. 8:**
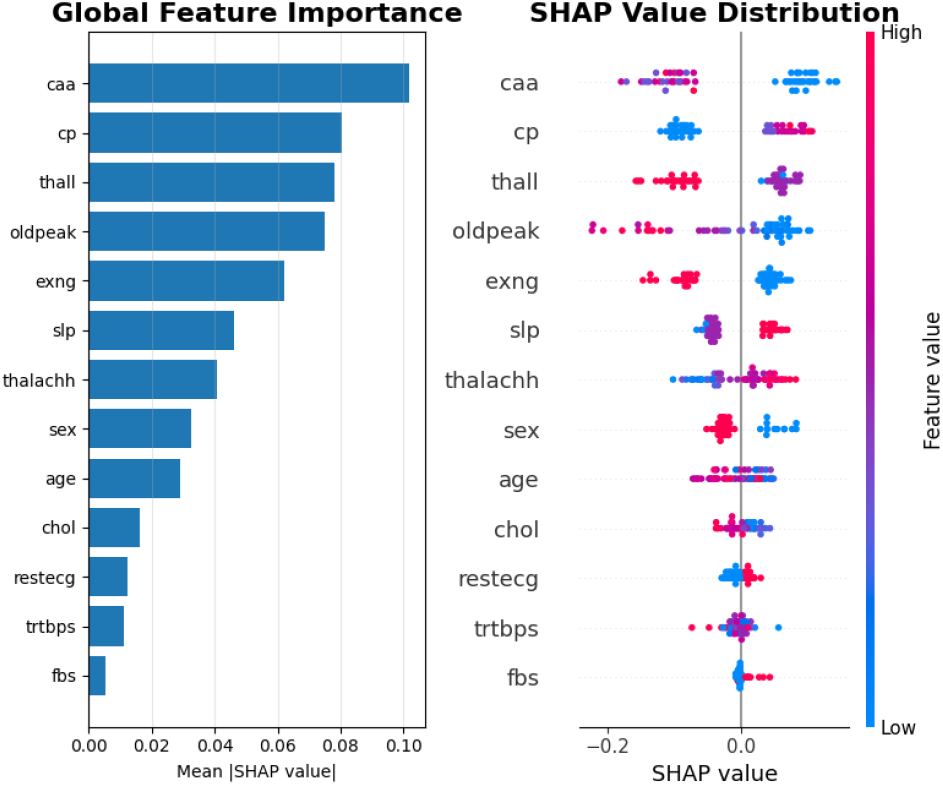
Global Feature Importance and SHAP Value Distribution for Dataset 2.

**Fig. 9:**
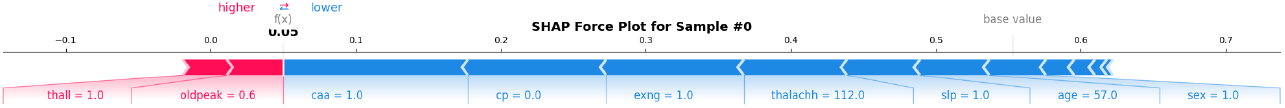
SHAP Force Plot for Sample 0 in Dataset 2.

These SHAP analyses provide insight into the most influential clinical features for heart failure prediction, enhancing model transparency and supporting interpretable clinical decision-making.

## 5 Discussion

The results demonstrated that model performance was strongly influenced by the characteristics of the data. The highest scores were achieved by the MLP model on the first two datasets which is shown in Table 13. For Dataset 1, an accuracy of 0.864 was recorded, along with a recall of 0.859, a precision of 0.903, an ROC-AUC of 0.865, and a PR-AUC of 0.957. Similarly, for Dataset 2, scores of 0.836 (accuracy), 0.812 (recall), 0.864 (precision), 0.837 (ROC-AUC), and 0.906 (PR-AUC) were obtained.

The model’s performance was further validated by its confusion matrices which is shown in Fig. 5. For Dataset 1, a high number of correct predictions was demonstrated, with 92 true positives (TP) and 67 true negatives (TN) being identified. Comparatively few errors were made, with only 15 false negatives (FN) and 10 false positives (FP) being recorded. A similar trend was shown in Dataset 2, where the error profile suggested a conservative model: a very low number of false alarms was produced (FP = 4), while the number of missed positive cases was also kept low (FN = 6).

Furthermore, the SHAP analysis provided critical insights into the feature importance patterns across different datasets. For Dataset 1, the global feature importance plot (Fig. 6) revealed that clinical indicators such as *ST Slope* and *ExerciseAngina* were the most influential predictors, suggesting strong physiological signals in well balanced data. In contrast, the analysis for Dataset 2 (Fig. 8) showed a more distributed feature importance pattern, with factors like *Coronary artery aneurysm (CAA)* and *Constrictive pericarditis (CP)* gaining relative prominence, potentially indicating that the model relied on different feature combinations when faced with varying data characteristics. The SHAP force plot for Dataset 2 (Fig. 9) further demonstrated how these features interact at the individual prediction level, revealing the additive contributions that drive specific clinical outcomes. This variation in feature importance across datasets underscores the context-dependent nature of predictive models and highlights the need for dataset-specific interpretability analysis to ensure clinically meaningful explanations.

Furthermore, the findings of this study have significant implications for hospital mortality prediction. By systematically evaluating multiple ML models using cross-validated approaches, we show that model performance is highly influenced by the characteristics and balance of the underlying data. This highlights the necessity of comparative benchmarking across diverse models, rather than relying on a single algorithm, to identify the most reliable predictor in a specific clinical context. In hospital mortality prediction, where false negatives carry substantial risk, cross-validated comparisons provide a robust framework for selecting models that optimize both sensitivity and generalizability. Consequently, the methodology presented here in can serve as a foundation for developing clinically deployable decision-support tools, enabling improved patient stratification, early risk detection, and targeted interventions in critical care settings.

## 6 Conclusion

This study systematically evaluated multiple ML models across two datasets to predict heart failure, highlighting both their potential and limitations. The results demonstrated that model performance is highly dependent on the characteristics and balance of the underlying data. The MLP model achieved the highest performance on Dataset 1 and Dataset 2. Importantly, the incorporation of SHapley Additive exPlanations (SHAP) provided critical interpretability, revealing the context dependent nature of feature importance. For instance, clinical indicators such as *ST Slope* and *ExerciseAngina* were most influential in Dataset 1, whereas *Coronary artery aneurysm (CAA)* and *Constrictive pericarditis (CP)* gained prominence in Dataset 2. This underscores the need for dataset-specific interpretability analyses to provide clinically meaningful insights. Overall, the findings suggest that while ML models can achieve strong predictive performance in well-balanced datasets, challenges remain in handling unbalanced data and ensuring robust generalization. Future work should focus on improving recall to minimize false negatives in critical healthcare applications, through strategies such as class rebalancing, advanced feature engineering, and hyperparameter optimization.

## Data Availability

The data used in this study are publicly available on Kaggle.
https://doi.org/10.34740/KAGGLE/DS/2162210
https://doi.org/10.34740/KAGGLE/DS/678821

https://doi.org/10.34740/KAGGLE/DS/678821

## Declarations

### Ethics approval

Not applicable (this study used publicly available, de-identified datasets with no human subject interaction).

### Consent for participation and publication

Not applicable.

### Availability of data and materials

The datasets analyzed during this study are publicly available on Kaggle: (1) Fedesoriano: *Heart Failure Prediction Dataset* (2021), DOI: 10.34740/KAGGLE/DS/2162210; and (2) Rashikrahmanpritom: *Heart Attack Analysis and Prediction Dataset* (2021), DOI: 10.34740/KAGGLE/DS/678821. Both datasets are open-access and available to the public through the Kaggle platform.

### Code availability

No custom code was developed or released as part of this study. The analysis was conducted using standard open-source Python libraries (e.g., pandas, scikit-learn, xgboost).

### Competing interests

The authors declare no competing interests. None of the authors is a member of the Editorial Board of this journal.

### Funding

This research did not receive any specific grant from funding agencies in the public, commercial, or not-for-profit sectors.

### Authors’ contributions

MSA: Conceptualization, methodology, model implementation, manuscript drafting. TI: Data preprocessing, statistical analysis, manuscript review and editing. Both authors read and approved the final manuscript.

## Acknowledgments

The authors thank the Information & Computer Science Department, King Fahd University of Petroleum and Minerals (KFUPM), for academic support.

## Clinical Trial Registration

Not applicable.

## Permissions

All figures and tables are original to this work. No copyrighted or previously published materials were reused.

## Notes

### Competing Interest Statement

The authors have declared no competing interest.

### Funding Statement

This study did not receive any external funding.

### Author Declarations

The study used (or will use) ONLY openly available human data that were originally located at: https://doi.org/10.34740/KAGGLE/DS/2162210 https://doi.org/10.34740/KAGGLE/DS/678821

### Summary of Updates

In this revised version of the manuscript, we have corrected an inconsistency in the data description. The original abstract incorrectly stated that three datasets were used for model development, whereas only two publicly available datasets from Kaggle were utilized: the Heart Failure Prediction Dataset and the Heart Attack Analysis and Prediction Dataset. This has now been clarified throughout the manuscript to ensure accuracy and consistency. In addition, the Results section has been refined to present a more balanced and comprehensive analysis of model performance. These updates improve the clarity, reproducibility, and overall quality of the manuscript without altering the main conclusions.

